# DWI-ADC Mismatch Predicts Infarct Growth and Endovascular Thrombectomy Outcomes in Anterior Circulation Stroke

**DOI:** 10.1101/2025.03.13.25323930

**Authors:** Chen-Hua Chiang, Chia-Chuan Liu, Chi-Lun Weng, Lung Chan, Lillian Tsao, David Yen-Ting Chen

## Abstract

**Background:** Current automatic software uses a fixed apparent diffusion coefficient (ADC) threshold (≤620×10⁻⁶ mm²/s) to quantify stroke volume for endovascular thrombectomy (EVT) decision-making. In contrast, clinical evaluation relies on visual identification of DWI hyperintensity with corresponding ADC hypointensity—which may exhibit varied ADC values in hyperacute stroke. This study investigates the clinical and imaging significance of the significant discrepancy between DWI hyperintense volume and ADC ≤ 620 × 10⁻⁶ mm²/s volume on pre-EVT magnetic resonance imaging (MRI), defined as DWI-ADC mismatch.

**Method:** This retrospective, single-center study analyzed consecutive patients diagnosed with acute ischemic stroke (AIS) between January 2018 and January 2020. Inclusion criteria consisted of patients with symptomatic anterior circulation large vessel occlusion (LVO) within 24 hours and treated by EVT, with high-quality pre- and post-EVT MRI scans. Infarct segmentation was performed by neuroradiologists. DWI-ADC mismatch was defined as a ratio of segmented DWI volume to ADC volume (≤620×10⁻⁶ mm²/s) ≥2. Data on demographics, baseline and follow-up clinical and imaging characteristics, procedural details, and outcomes were collected and analyzed.

**Result:** 73 patients were included in the study, with 20 patients (27.4%) demonstrating a DWI-ADC mismatch. The mismatch group exhibited a significantly slower infarct growth rate (3.8 vs. 7.5 ml/h, *P* = 0.04), higher prevalence of parent artery stenosis (65% vs. 20.8%, *P* < 0.001) and a greater need for angioplasty and/or stenting (50% vs. 17%, *P* < 0.001). Imaging analysis showed a higher percentage of DWI reversal (37.7% vs. 21.2%, *P* = 0.02) and a trend toward lower ADC lowering percentage (25.3% vs. 32.2%, *P* = 0.05) in the mismatch group. Demographics, baseline and follow-up lesion volumes, and functional outcomes were comparable between groups.

**Conclusion:** DWI-ADC mismatch identifies a distinct subgroup of stroke patients with slower infarct progression and unique procedural needs. Incorporating mismatch evaluation into pre-EVT imaging may refine patient selection and optimize treatment strategies.

## Introduction

Ischemic infarct volume quantification is critical for guiding EVT decisions, both in determining patient eligibility pre-EVT ^1,2^ and predicting outcomes post-EVT.^3,4^ Current automatic software uses an apparent diffusion coefficient (ADC) threshold of ≤ 620 × 10⁻⁶ mm²/s to identify and quantify irreversible infarct core volume.^5^ MRI with DWI and ADC sequences is the gold standard tool for ischemic infarct detection.^6,7^

Clinicians often rely on visually identifying DWI hyperintense lesions with corresponding ADC hypointensity to detect ischemic infarcts. However, in hyperacute strokes, these ADC hypointense lesions frequently encompass a range of ADC values, reflecting varying tissue viability. Using visible DWI lesions to estimate infarct core has also been challenged, although DWI lesion is generally a reliable signature for infarct core.^8^ Research shows that DWI lesions may reverse after successful reperfusion with EVT or thrombolysis, particularly when reperfusion is achieved rapidly. ^9–12^ This suggests that relying on DWI lesions alone may overlook tissue salvage potential.

The limitations of both fixed ADC thresholds and visual DWI detection introduce risks of suboptimal clinical decision-making. Variations in ADC thresholds have been shown to produce discrepancies in infarct volume measurements,^13^ potentially leading to the exclusion of patients who could benefit from EVT or the inclusion of those who might not. This variability underscores the need for a more nuanced approach to ischemic core assessment.

Our study addresses this gap by investigating the prevalence and clinical significance of DWI-ADC mismatch, defined as significant differences between DWI hyperintense volume and ADC ≤ 620 × 10⁻⁶ mm²/s volume. By quantitatively assessing these lesions pre- and post-EVT, we aim to evaluate their evolution and implications for clinical decision-making. We hypothesize that DWI hyperintensity with ADC hypointensity but ADC values > 620 × 10⁻⁶ mm²/s reflects tissue with uncertain viability. Understanding these discrepancies may enhance infarct characterization, improve patient selection, and optimize EVT outcomes.

## Method

### Study Design

This retrospective, single-center study included consecutive acute ischemic stroke patients (AIS) patients between January 2018 and January 2020, fulfilling the following criteria: (1) first presentation of symptomatic anterior circulation large vessel occlusion (LVO): defined as occlusion mainly involved the internal carotid artery, M1 or M2 segments of middle cerebral artery), (2) documented time from symptom onset (or last seen well for patients with unknown symptom onset or wake-up stroke) to EVT triage within 24 hours, (3) underwent EVT, (4) available good-quality pretreatment MRI with DWI and ADC maps and (5) available good-quality follow-up MRI within 5 days of EVT.

This study was approved by the institutional review boards from our institution. Informed consent was waived by the institutional review boards due to the retrospective design of the study.

### Image Acquisition

MR images were performed using two machines, including 1.5T (Signa HDX, GE Healthcare, Milwaukee, WI, USA) and 3T scanners (Discovery MR750, GE Healthcare, Milwaukee, WI, USA). The parameters for the DWI sequences for the 1.5 T MRI included the following: repetition time = 5336 milliseconds; echo time = minimum; image matrix size = 160*120; interslice gap = 2 mm; number of slices = 20 - 24; slice thickness = 5 mm; b-value = 1000 s/mm2. For the 3 T MRI, the parameters were as follows: repetition time: 8000 milliseconds; echo time = minimum; image matrix: 128*128. FOV was 230 mm, section thickness was 5 mm and interslice gap was 2 mm for both scanners.

### Baseline and Final Infarct Segmentation

Visual DWI hyperintense infarcts with corresponding ADC hypointensity were segmented by neuroradiologists (C.H.C. and C.L.W.) using Medical Imaging Interaction Toolkit (MITK) ^14,15^ in the pre-treatment MRI on DWI sequence. The neuroradiologists were blinded to clinical and outcome data during segmentation to minimize bias. Comparison to T2 FLAIR images or ADC b=0 images were done to exclude T2 shine-through effect caused by the possibility of edema, or infarct mimics such as non-specific white matter change, old insult or even artifact. For cases that were still difficult to interpret, a consensus between two radiologists was achieved.

For final infarct segmentation, same method was employed. Presence of hemorrhagic transformation and infarcts extending to the contralateral hemisphere were included as final infarct to avoid underestimation of final infarct volume. ^16^

### Calculation and definition of DWI-ADC mismatch

The segmented DWI lesions were saved into Neuroimaging Informatics Technology Initiative (NIFTI) files. Direct extraction of ADC values within the segmented DWI lesions was performed in a voxel-based analysis using Python. Individual volume of ADC ≤ 620 and > 620 ×10^−6^ mm^2^/s within the segmented DWI lesion were identified and recorded. The DWI-ADC volume ratio was calculated by dividing the segmented DWI volume over ADC ≤ 620 (×10^−6^ mm^2^/s) volume. DWI-ADC mismatch was defined as a ratio ≥ 2, and no mismatch when the ratio was < 2.

### Clinical Data Collection

The patient demographics at baseline were obtained from the institutional EVT registry or electronic medical records. For all patients, we recorded the time from the first symptom recognition or last known well time (LKWT) to the beginning of MRI time. Infarct growth rate (IGR) was calculated by this formula: IGR = Baseline DWI lesion volume ÷ Time from last known well to imaging (in decimal hours).

### Analysis of DWI lesion evolution

Baseline DWI lesion masks were co-registered to follow-up MRIs using the FMRIB Software Library (FSL) for voxel-wise comparison of DWI lesion evolution.^17,18^ The transformation matrix derived from DWI b=0 images was applied to pre-EVT lesion masks to generate co-registered masks, minimizing inter-scan errors. Overlaid DWI-positive areas were visually inspected by a neuroradiologist to ensure quality.

The following volumes and percentage of volumes were calculated. The percentage of volumes were calculated to account for variability in the baseline lesions size ^11^:

- DWI lesion reversal (DWIR): Defined as the volume of voxels returning to normal on follow-up DWI. Calculated as: Percentage of DWIR (DWIR%) = (DWIR volume ÷ baseline DWI hyperintense volume) × 100.
- DWI lesion growth (DWIG): Defined as new hyperintense voxels on follow-up DWI. Calculated as: Percentage of DWIG (DWIG%) = (DWIG volume ÷ baseline DWI hyperintense volume) × 100.
- DWI unchanged (DWIU): Defined as baseline hyperintense voxels remaining unchanged on follow-up DWI. Calculated as: Percentage of DWI unchanged volume (DWIU%) = (DWIU ÷ baseline DWI hyperintense volume) × 100.

### Analysis of ADC lesion evolution

Changes in ADC lesion volumes (≤ 620 and > 620 ×10^−6^ mm^2^/s) were analyzed by co-registering pre-EVT DWI and ADC images to post-EVT DWI images.

Transformation matrices from DWI b=0 images were applied consistently to align DWI b=1000, ADC, and lesion masks, ensuring precise voxel-wise evaluation. The following volumes and percentage of volumes were then calculated:

- ADC lesion reversal (ADCR): Defined as voxels with ADC ≤ 620 × 10⁻⁶ mm²/s on baseline images that reverted to ADC > 620 × 10⁻⁶ mm²/s on follow-up images. Calculated as: ADCR% = (ADCR volume ÷ baseline ADC ≤ 620 × 10⁻⁶ mm²/s volume) × 100.
- ADC irreversible (ADCI): Defined as voxels with ADC ≤ 620 × 10⁻⁶ mm²/s on baseline images that remain ≤ 620 × 10⁻⁶ mm²/s on follow-up images. Calculated as: ADCI% = (ADCI volume ÷ baseline ADC ≤ 620 × 10⁻⁶ mm²/s volume) × 100.
- ADC lowering (ADCL): Defined as voxels with ADC > 620 × 10⁻⁶ mm²/s on baseline images that changed to ADC ≤ 620 × 10⁻⁶ mm²/s on follow-up images. Calculated as: ADCL% = (ADCL volume ÷ baseline ADC > 620 × 10⁻⁶ mm²/s volume) × 100.
- ADC unchanged (ADCU): Defined as voxels with ADC > 620 × 10⁻⁶ mm²/s on baseline images that remained > 620 × 10⁻⁶ mm²/s on follow-up images. Calculated as: ADCU% = (ADCU volume ÷ baseline ADC > 620 × 10⁻⁶ mm²/s volume) × 100.

### Evaluation of Endovascular Recanalization and procedural variables

Successful vessel recanalization status was defined as modified Thrombolysis in Cerebral Infarction grade of ≥2b on angiographic images.^19^ Definition of parent artery stenosis (PAS) is the presence of stenosis at the occluded artery or artery proximal to the occluded artery. PAS or other procedural variables such as performance of angioplasty or stenting during the procedure were collected from electronic medical records (EMR) or radiology information system (RIS). If the record was unclear or not available, then their images were reviewed and evaluated by neuroradiologist (C.H.C).

### Assessment of Outcome After Endovascular Treatment

Intracranial hemorrhages after EVT were classified on follow-up imaging according to the Heidelberg Bleeding Classification criteria. ^20^ The classification of reperfusion hemorrhages was performed by neuroradiologist (C.H.C). Ninety-day modified Rankin Scale (mRS) were defined as the primary outcome measure. 90-day mRS scores of 0 to 2 were considered as good clinical outcomes. The National Institutes of Health Stroke Scale score (NIHSS) was used to evaluate baseline stroke severity and discharge outcome. Both the mRS scores and NIHSS were determined by a stroke neurologist or registered study nurse.

### Statistical Analysis

Patient demographics, medical history, and imaging findings were compared between mismatched and no mismatched patients. Categorical variables were reported as counts and percentages. Continuous variables were reported as mean and SD or as median and interquartile range. Categorical variables were compared between groups using chi-squared test (χ 2) or Fisher exact test when appropriate. Continuous variables were compared using Mann-Whitney U test. Cases with missing values were excluded from the respective analyses. A 2-tailed P of <0.05 was considered significant for all statistical tests. All P smaller than 0.001 were reported as P<0.001.

## Result

### Baseline Patient Characteristics

A total of 73 patients met the inclusion criteria. Figure 1 shows the flowchart of eligible patients. Among these patients, the average age was 72.8 years and 36 were men (49.3%). Of these patients, 20 patients (27.4%) reached our definition of DWI-ADC mismatch (Figure 2). The baseline NIHSS and stroke etiology between the mismatch and no mismatch group showed no statistically significant difference. The characteristics of patients’ variables are summarized in the Table 1.

**Figure 1.**
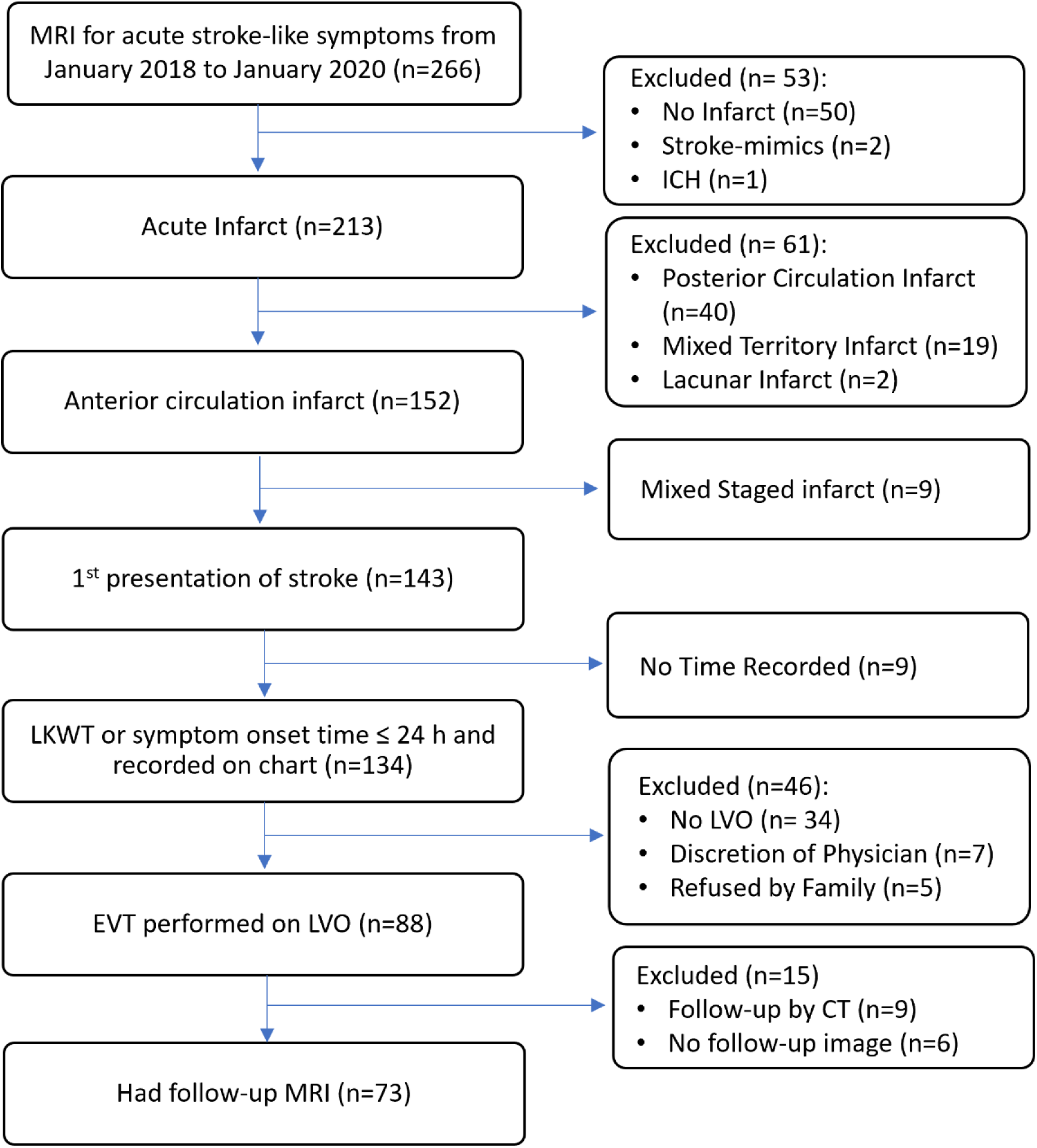
Study flowchart illustrating patient inclusion criteria.

**Figure 2.**
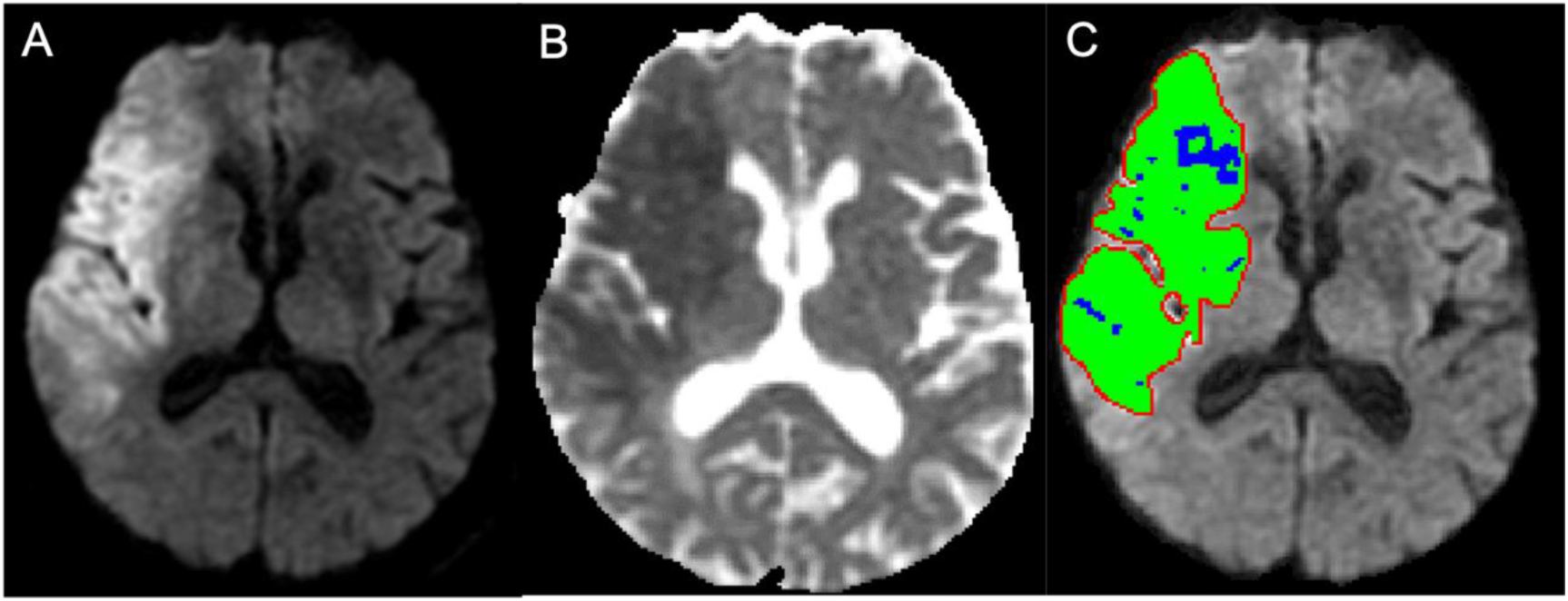
Example of DWI-ADC mismatch. A, DWI of a patient with right middle cerebral artery infarction. B, Corresponding ADC map. C, Segmentation mask (red line); blue represents ADC ≤ 620 ×10−6 mm²/s, green represents area that is ADC > 620 ×10−6 mm²/s. The DWI/ADC ratio: 13.9 for this slice, 2.2 for this case overall, indicating a significant mismatch between DWI and ADC volumes.

**Table 1.** Baseline Patient, Clinical, Procedural and Outcome Characteristics Stratified by DWI-ADC Mismatch.

|  | All patients<br>(n= 73) | DWI-ADC mismatch<br>(n= 20) | No DWI-ADC mismatch<br>(n=53) | *P* value<sup>‡</sup> |
| --- | --- | --- | --- | --- |
| Patient Characteristics |  |  |  |  |
| Age, y; mean (SD) | 72.8<br>(10.8) | 74.7 (9.0) | 72.1 (11.3) | 0.430 |
| Male sex, n (%) | 36 (49.3) | 10 (50) | 26 (49.1) | 0.943 |
| Baseline NIHSS, mean (SD) | 17.7<br>(5.9) | 16.3 (6.5) | 18.2 (5.6) | 0.280 |
| TOAST classification (missing=5) |  |  |  | 0.183 |
| LAA | 30 (44.1) | 9 (52.9) | 21 (41.2) |  |
| Cardioembolism | 34 (50) | 8 (47.1) | 26 (51.0) |  |
| Small-vessel/Other/Undetermined | 4 (5.9) | 0 (0) | 4 (7.8) |  |
| Clinical Variables |  |  |  |  |
| Onset to image time, Decimal hour; median (IQR) | 2.9 (3.1) | 3.6 (3.0) | 2.8 (2.8) | 0.164 |
| Infarct growth rate, ml/h; median (IQR) | 5.7 (15.3) | 3.8 (8.8) | 7.5 (15.0) | 0.040 |
| Intravenous tPA | 31 (42.5) | 11 (55) | 20 (37.7) | 0.183 |
| Follow-up MRI from EVT, Days; median (IQR) | 1 (1) | 1.5 (1) | 1 (1) | 0.936 |
| **Procedural Variables** |  |  |  |  |
| Occlusion site<sup>*</sup> |  |  |  | 0.678 |
| ICA; n (%) | 17 (23.3) | 4 (20.0) | 13 (24.5) |  |
| M1; n (%) | 42 (57.5) | 11 (55.0) | 31 (58.5) |  |
| M2; n (%) | 29 (39.7) | 11 (55.0) | 18 (34.0) |  |
| Other arteries<sup>#</sup>; n (%) | 3 (4.1) | 1 (5.0) | 2 (3.8) |  |
| Presence of parent artery stenosis; n (%) | 24 (32.9) | 13 (65.0) | 11 (20.8) | <0.001 |
| Angioplasty and/or stenting; n (%) | 19 (26.0) | 10 (50.0) | 9 (17.0) | <0.001 |
| **Outcome Variables** |  |  |  |  |
| mTICI score≥2b, n (%) | 69 (94.5) | 20 (100.0) | 49 (92.5) | 0.570 |
| Midline shift, n (%) | 5 (6.9) | 0 (0) | 5 (9.4) | 0.314 |
| Discharge NIHSS, median (IQR)<sup>†</sup> | 11 (11.0) | 12 (11.5) | 13 (14.8) | 0.562 |
| Discharge mRS, median (IQR) | 4 (2.0) | 4 (1.3) | 4 (2.0) | 0.238 |
| 3-month mRS, median (IQR) | 4 (1.0) | 3.5 (2.0) | 4 (2.0) | 0.276 |
| 3-month mRS 0-2, n (%) | 18 (24.7) | 7 (35.0) | 11 (20.8) | 0.208 |
| **Hemorrhagic Transformation** |  |  |  |  |
| No Hemorrhage | 27 (37.0) | 10 (50.0) | 17 (32.1) | 0.174 |
| Any Hemorrhage, n (%)<sup>§</sup> | 45/72<br>(62.5) | 10 (50.0) | 35/52 (67.3) | 0.174 |
| HI1 | 9 (12.3) | 3 (15) | 6 (11.3) |  |
| HI2 | 7 (9.6) | 1 (5) | 6 (11.3) |  |
| PH1 | 7 (9.6) | 1 (5) | 6 (11.3) |  |
| PH2 | 20 (27.4) | 5 (25) | 15 (28.3) |  |
| Class 3 hemorrhage<sup>§§</sup> | 5 (6.8) | 1 (5) | 4 (7.3) |  |
| Hemorrhage other than HT, n (%) | 25/72<br>(34.7) | 6 (30.0) | 19/52 (28.8) | 0.602 |

<sup>¥</sup> P value is calculated with either Mann-Whitney U test for continuous variables or a Chi-square test or Fisher-exact test for counts for one cell ≤5 for categorical variables
\* Occlusion site was determined by EVT findings and may involve multiple arteries (e.g., ICA + M1), resulting in summation percentages exceeding 100% for some cases.

### Clinical and Procedural Characteristics

The onset to image time were comparable between the mismatch and no mismatch group (3.61 hours versus 2.79 hours), however, there is statistically significant IGR difference between mismatch and no mismatch group (3.75 ml/h versus 7.49 ml/h, *P*=0.04). There is no difference between mismatch and no mismatch group in terms of occlusion site, however, the mismatch group were more likely to have parent artery stenosis (65% versus 20.8%, *P*<0.001) and required to have angioplasty and/or stenting (50% versus 17%, *P*<0.001) during EVT (Table 1).

### Outcome Variables According to DWI-ADC Mismatch

The overall mTICI scores ≥2b were 94.5% and were comparable across groups (*P*=0.57). No difference across groups in terms of percentage of post-EVT hemorrhage (*P*=0.174). There were also no differences in terms of discharge NIHSS, mRS, 3-month mRS and proportion of good functional outcome at 3-month (35% versus 20.8%, *P*=0.208) (Table 1).

### Quantitative Evaluation of Image Variables According to DWI-ADC Mismatch

The baseline DWI lesion volume was smaller in the mismatched group compared to the no mismatch group (11.5 ml vs. 20.5 ml, *P*=0.17), but this difference was not statistically significant. Similarly, follow-up DWI lesion volumes (38.8 ml vs. 44.7 ml, *P*=0.741) and ADC ≤ 620 (×10^−6^ mm^2^/s) lesion volumes (15.5 ml vs. 18.5 ml, *P*=0.542) showed no significant differences between groups. However, the mismatch group had a higher DWIR% (37.7% vs. 21.2%, *P*=0.023), despite no difference in DWIR volume (6.4 ml vs. 5.8 ml, *P*=0.826). DWIU% was lower in the mismatch group (62.4% vs. 78.8%, *P*=0.026), though DWIU volume did not differ significantly (5.0 ml vs. 14.7 ml, *P*=0.767). No significant differences were observed for DWIG volume or percentage (Table 2). Figure 3 illustrates DWI reversal in a patient with DWI-ADC mismatch.

**Figure 3.**
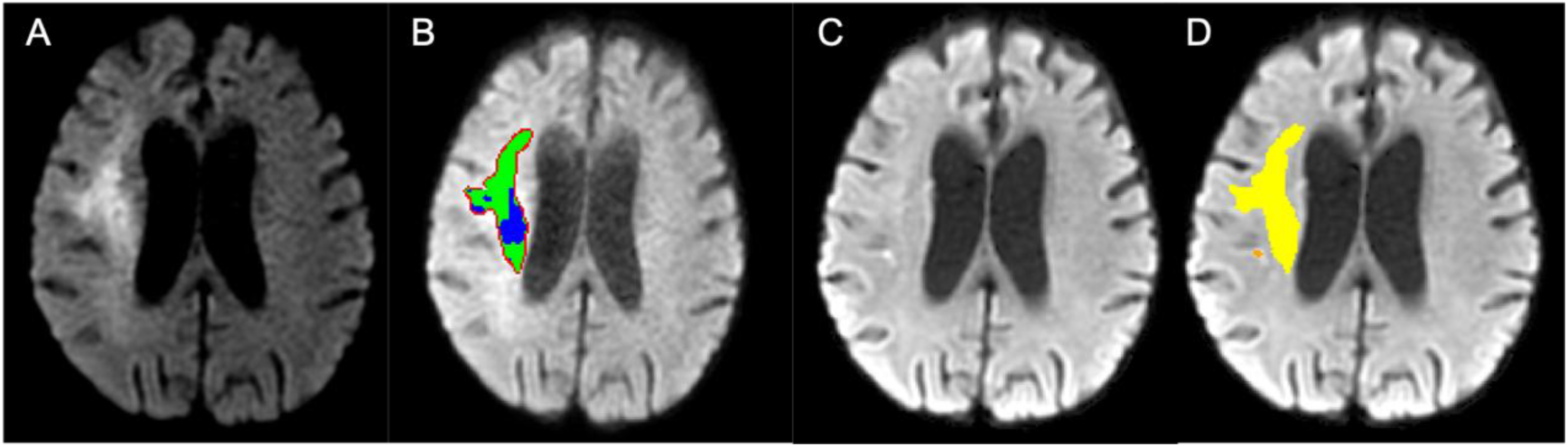
Example of DWI-ADC mismatch with DWI lesion evolution. A, Baseline DWI showing right middle cerebral artery infarction. B, DWI-ADC mismatch with color coding as in Figure 1 (DWI/ADC ratio: 3.0 for this slice). C, Follow-up DWI. D, Follow-up DWI with yellow indicating DWI reversal and orange indicating DWI growth.

**Table 2.** Quantitative Evaluation of Image Variables According to DWI-ADC Mismatch.

|  | All patients (n= 73) | DWI-ADC mismatch (n= 20) | No DWI-ADC mismatch (n= 53) | P value |
| --- | --- | --- | --- | --- |
| **Baseline Imaging Variables** |  |  |  |  |
| Baseline DWI lesion volume, ml; median (IQR) | 19.8 (55.2) | 11.5 (30.6) | 20.9 (64.7) | 0.171 |
| Baseline ≤ ADC 620 volume, ml; median (IQR) | 12.1 (39.0) | 3.25 (14.1) | 13.5 (41.1) | 0.011 |
| Baseline ADC > 620 volume, ml; median (IQR) | 7.5 (19.7) | 8.6 (21.4) | 7.3 (16.4) | 0.928 |
| Mismatch ratio; median (IQR) | 1.62 (0.64) | 2.93 (1.53) | 1.49 (0.33) | <0.001 |
| **Follow-up Image Variables** |  |  |  |  |
| Follow-up DWI lesion volume, ml; median (IQR) | 39.6 (137.7) | 38.8 (167.1) | 44.7 (126.7) | 0.741 |
| Follow-up ADC ≤ 620 volume, ml; median (IQR) | 17.1 (46.8) | 15.5 (81.7) | 18.5 (38.5) | 0.542 |
| Follow-up ADC > 620 volume, ml; median (IQR) | 21.3 (41.2) | 19.42 (42.3) | 21.54 (39.6) | 0.726 |
| **DWI Lesion Evolution** |  |  |  |  |
| DWIR, ml; median (IQR) | 6.1 (11.1) | 6.4 (11.0) | 5.8 (10.7) | 0.826 |
| DWIR%, %; median (IQR) | 25.6 (29.5) | 37.7 (42.0) | 21.2 (23.2) | 0.023 |
| DWIG, ml; median (IQR) | 25.2 (49.7) | 20.1 (51.0) | 25.3 (50.0) | 0.889 |
| DWIG%, %; median (IQR) | 90.7 (146.2) | 93.7 (323.3) | 90.7 (135.6) | 0.682 |
| DWIU, ml; median (IQR) | 13.0 (47.0) | 4.9 (23.3) | 14.7 (53.2) | 0.767 |
| DWIU%, %; median (IQR) | 74.4 (29.5) | 62.4 (42.3) | 78.8 (23.2) | 0.026 |
| **ADC Lesion Evolution** |  |  |  |  |
| ADCR, ml; median (IQR) | 5.1 (12.8) | 1.6 (8.7) | 6.2 (10.4) | 0.027 |
| ADCR%, %; median (IQR) | 54.2 (44.3) | 52.7 (38.1) | 54.4 (33.4) | 0.529 |
| ADCL, ml; median (IQR) | 2.1 (6.8) | 1.8 (5.4) | 2.2 (10.9) | 0.337 |
| ADCL%, %; median (IQR) | 28.7 (28.7) | 25.3 (13.7) | 32.2 (30.0) | 0.050 |
| ADCI, ml; median (IQR) | 3.9 (11.8) | 1.2 (3.6) | 6.2 (13.9) | 0.003 |
| ADCI%, %; median (IQR) | 45.6 (43.8) | 44.2 (36.1) | 45.5 (44.3) | 0.184 |
| ADCU, ml; median (IQR) | 7.4 (16.8) | 7.7 (17.8) | 7.0 (14.0) | 0.741 |
| ADCU%, %; median (IQR) | 71.6 (28.7) | 74.7 (13.2) | 67.5 (30.1) | 0.051 |

For ADC lesion evolution, a significant difference was observed in ADCR volume (1.6 ml vs. 6.2 ml, *P*=0.027), but not in ADCR% (52.7% vs. 54.4%, *P*=0.529). Similarly, while ADCI volume showed significant differences (1.2 ml vs. 6.3 ml, *P*=0.337), ADCI% did not (44.2% vs. 45.6%, *P*=0.184). No significant differences were found in ADCL volume (1.8 ml vs. 2.2 ml, *P*=0.337), although a borderline significant difference was noted in ADCL% (25.3% vs. 32.2%, *P*=0.050). ADCU volume was similar between groups (7.7 ml vs. 7.0 ml, P=0.741), but ADCU% showed borderline significance (74.7% vs. 67.5%, *P*=0.051). Figure 4 depicts an example of DWI-ADC mismatch with ADC lesion evolution.

**Figure 4.**
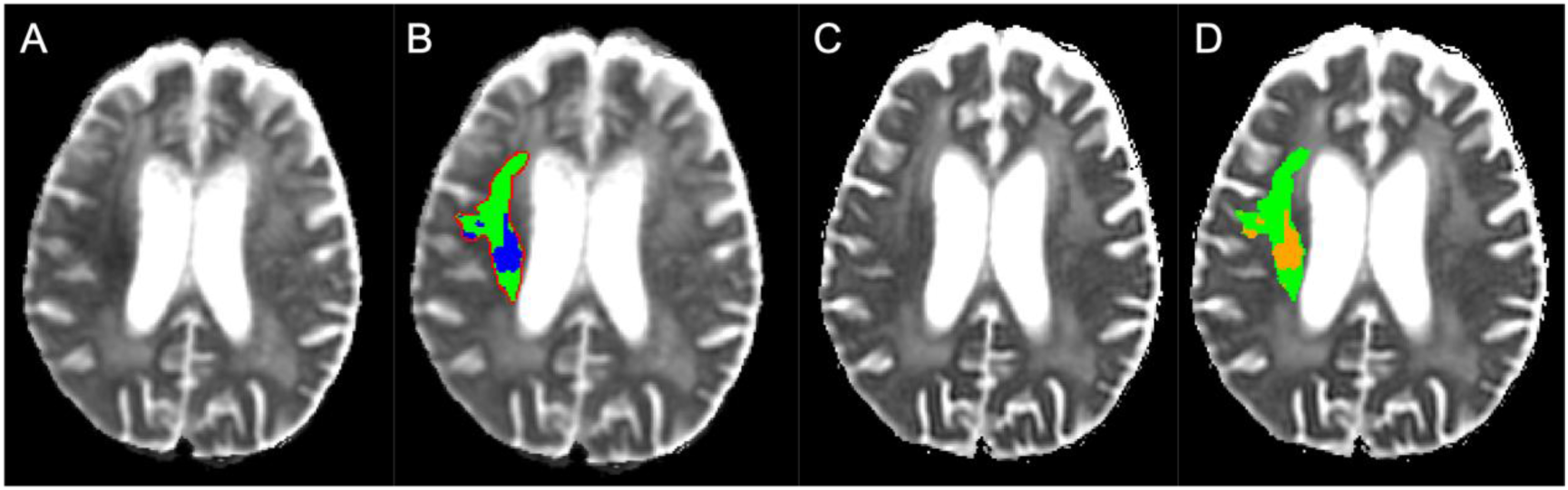
Example of DWI-ADC mismatch with ADC lesion evolution. Same slice as Figure 3. A, Baseline ADC map. B, DWI-ADC mismatch (color coding as in Figure 1) overlaid on ADC map. C, Follow-up ADC map. D, Follow-up ADC map with orange indicating ADC reversal and green indicating ADC unchanged.

## Discussion

In our study, 27.4% of patients met the definition of DWI-ADC mismatch (pre-EVT DWI lesion volume/ADC ≤ 620 (×10⁻⁶ mm²/s) lesion volume ≥ 2). These patients exhibited significantly lower infarct growth rates and a higher prevalence of parent artery stenosis, necessitating more frequent angioplasty and/or stenting during EVT. DWI lesion analysis revealed a significantly higher percentage of DWI lesion reversal (DWIR%) in the mismatch group, alongside a lower ADCL% that approached statistical significance. These findings suggest that DWI-ADC mismatch may identify a subset of patients with slower infarct progression and greater potential for complex vascular interventions. Additionally, successful reperfusion achieved through EVT may be associated with greater infarct volume recovery in cases of DWI-ADC mismatch.

ADC thresholds have been widely used for ischemic core quantification. Purushotham et al. established an ADC threshold of ≤ 620 × 10⁻⁶ mm²/s, commonly adopted in automated software, though its sensitivity (69%) and specificity (78%) remain moderate.^5^ Umemura et al. proposed a lower threshold of 520 × 10⁻⁶ mm²/s to avoid DWI reversal, achieving slightly better sensitivity and specificity of 71% and 83%, respectively. ^21^ However, ADC values are dynamic and evolve with time and perfusion changes during ongoing infarction.^22,23^ Our findings emphasize the limitations of relying on static ADC thresholds and reinforces that DWI hyperintensity with ADC hypointensity may indicate tissue with uncertain viability. Notably, 25.3–32.2% of baseline ADC > 620 ×10⁻⁶ mm²/s showed further ADC reduction post-EVT despite high reperfusion rates, while 52.7–54.4% of baseline ADC ≤ 620 × 10⁻⁶ mm²/s region showed reversal. These findings underscore the limitations of current ADC-based ischemic core definitions and aligns with Goyal et al.’s concept of “severely ischemic tissue with uncertain viability (SIT-uv).”^6^. Larger, multi-center studies are essential to validate these findings and refine treatment strategies.

Collateral circulation may contribute to the observed DWI-ADC mismatch phenomenon, which is a critical determinant of infarct dynamics and EVT outcomes. While DWI reflects ischemic stress, ADC values capture tissue viability, which depends on collateral supply. Although collateral status was not assessed directly in this study due to our reliance on MRI for pre-EVT evaluation, indirect markers such as parent artery stenosis and infarct growth rate suggest its influence. Chronic carotid stenosis has been shown to associate with better intracranial collateral pathwasy,^24^ while better collateral has been associated with smaller ischemic core^25^ and predict patient outcome.^26^ Moreover, good collateral status has been directly linked to slower early infarct growth rate, which itself predicts functional outcomes.^27,28^ This could explain why the mismatch group exhibited a higher prevalence of parent artery stenosis and slower infarct growth rates.

These observations may also explain the higher DWIR% observed in the mismatch group (37.7%) compared to the overall cohort rate (25.6%). This finding supports the hypothesis that more robust collateral supply in the mismatch group preserved tissue viability, increasing the likelihood of diffusion lesion reversal after successful reperfusion. Notably, the overall cohort rate is consistent with the 26.5% prevalence reported in a systematic review by Nagaraja et al., ^29^ adding reliability to our findings. The slower infarct growth rates observed in the mismatch group further support this hypothesis, as better collateral flow is known to maintain penumbral tissue viability.^27^ However, despite higher DWIR% and 100% successful reperfusion rate, only 35% of patients in the mismatch group achieved good 3-month functional outcomes. This discrepancy may be explained by the high baseline NIHSS (>14), which is associated with poor prognosis,^30^ and may reflect specific public health system factors, rehabilitation quality or unique characteristics of our patient population.^31^ This highlights that functional recovery at 3 months is multifactorial, and that DWIR% alone is insufficient to predict 3-month outcome.

The observed DWI-ADC mismatch can also be explained by the sensitivity of DWI signal intensity to variations in ADC values. DWI signal intensity follows the equation:

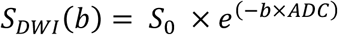

where b is the b-value, ADC is the apparent diffusion coefficient, SDWI(b) is signal intensity of the DWI at a specific b-value. ^32,33^ This exponential relationship means even slight ADC reductions result in substantial increases in DWI signal intensity, especially at higher b-values used for infarct detection. Additionally, T2 shine-through effects may contribute to lesion overestimation despite our best segmentation efforts. This dual influence—true diffusion restriction and potential T2 shine-through—can lead to DWI lesions appearing larger relative to ADC-defined volumes, contributing to the observed mismatch.

### Limitation

This study has several limitations. First, collateral status was not directly assessed as MRI currently lacks standardization and clinical validation for collateral assessment.^34^ Also although angiographic collateral grading remains the gold standard,^35^ the urgency of EVT often precludes its routine evaluation. Instead, we used indirect markers such as parent artery stenosis and infarct growth rate to infer collateral influence. Future studies incorporating advanced imaging modalities, such as multiphase CTA or perfusion MRI, are needed to validate these findings.

Second, the retrospective, single-center design and modest sample size may limit the generalizability of our results. Larger, multi-center studies are needed to confirm these observations and evaluate their applicability to broader populations.

Finally, the timing of follow-up imaging was not standardized across all cases, introducing potential variability in the measurement of infarct evolution. Nonetheless, given the heterogeneity in disease progression after EVT, a uniform MRI schedule would be impractical. While imaging within this timeframe generally minimizes the impact of early pseudonormalization of ADC,^36,37^ future studies with more uniform imaging protocols could enhance consistency and reliability.

## Conclusion

The presence of the presence of DWI-ADC mismatch is a frequent finding in AIS involving anterior circulation LVO prior to EVT. This mismatch is associated with slower infarct growth rates, higher rates of parent artery stenosis, and an increased need for angioplasty and/or stenting during EVT. Imaging analysis reveals that patients with DWI-ADC mismatch have significantly higher percentages of DWI reversal and borderline significant ADC lowering post-EVT. These findings highlight the importance of comprehensive imaging evaluations in AIS and suggest that identifying DWI-ADC mismatch may help define a distinct patient subgroup who could benefit from tailored therapeutic strategies in the future.

## Data Availability

All data supporting the findings of this study are available from the corresponding author upon reasonable request.

## Acknowledgments

The investigators thank the contribution from Hung Yi Chien and Ya-Ling Chang for their help on data collection and organization.

